# Effect of remedial teaching on academic performance of poorly performing students in Pharmacology

**DOI:** 10.1101/2021.04.13.21255387

**Authors:** Sophia B Modi, R Bindulatha Nair, GK Libu

## Abstract

**Purpose:** Significant learning difficulties requiring remediation has been observed to be experienced by many medical trainees. Research with regard to individualized remedial teaching based on pedagogical diagnosis is a strong need of the time. The objectives of this study were to assess the effect of remedial teaching in improving academic performance among poorly performing students in Pharmacology and to assess factors that could affect academic performance.

**Method:** The study was conducted in 2019. Academically poor performing students (< 50 % marks in Pharmacology first sessional exam) were selected after getting informed consent. After obtaining baseline information and *Study skills self-assessment inventory* information from all students, academically poor performing students (53 students) were identified and they were randomized into two groups. One group (26 students) received academic support alone. The second group (27 students) received academic support, sessions on study skills, stress-coping strategies and counselling regarding their academic and non-academic problems.

**Results:** The improvement in test scores among all participants of remedial sessions was statistically significant. Mean pre-test marks 5.27 ± 3.6 post test 14.63 ± 1.24 and the difference is statistically significant. Though apparently high 10.02 ± 3.25 Vs. 8.59 ± 3.55, the post intervention gain in scores is not statistically significant between Academic support + special package group Vs. Academic support alone group (P value 0.16)

**Conclusion:** Tailored or individualised remediation measures can greatly enhance the academic performance of undergraduate medical students and help them make satisfactory progress on the course.

## INTRODUCTION

The performance of undergraduate medical students in India and other developing countries is perceived to have largely declined.^1^Multiple stressors including academic burden, parental and peer pressure and even psychological ailments like depression, burn out and stress are seen more commonly among medical students. ^2^ Difficulty in understanding medium of instruction, sleep disorders and perceived parental and peer pressure and dissatisfaction with career choice have been significantly linked with poor performance.^3^

Remediation can be defined as “additional teaching above and beyond the standard curriculum, individualized to the learner who without the additional teaching would not achieve the necessary skills for the profession”. ^4^ Significant learning difficulties requiring remediation in the form of an individualized learning plan has been observed to be experienced by 7-28% of medical trainees ^5^,^6^.

The task of remediation to address these learning difficulties has been divided into four steps (SOAP model^7^), so as to guide teachers in approaching learners’ difficulties more effectively. These steps are 1) detecting problems, based on a subjective impression; 2) gathering and documenting objective data, according to diagnostic hypotheses; 3) making a pedagogical diagnosis based on this assessment; and 4) planning a targete^2^d remediation.

The subjective impressions, or intuitions, can be either shaped by direct observation of the learner in action or by interacting formally or informally with the student. Making a pedagogical diagnosis in medical education consists of “identifying discrepancy between the expected performance standard and the demonstrated performance, and then trying to establish the reason for underperformance”. ^8^

No standardized and universally accepted remediation processes have yet been developed. There is a strong need for further research with regard to remediation that is evidence-based, individualized, and targeted according to pedagogical diagnosis.

## RESEARCH QUESTION

Can remedial teaching improve academic performance among poorly performing students in Pharmacology?

## OBJECTIVE

1. To assess the effect of remedial teaching in improving academic performance among poorly performing students in Pharmacology as assessed by a pre-test and post-test
2. To assess factors that could affect academic performance.

## MATERIALS AND METHODS

**Study Design:** Quasi experimental

**Study setting:** Department of Pharmacology, Government Medical College, Kollam

**Study subjects:** 2^nd^ MBBS Students (currently having Pharmacology postings). All students who got less than 50 % marks in the first sessional exam of Pharmacology

**Study period**: July 2019 to August 2019 (2 months)

**Sample size:** All students who got less than 50% marks in the first sessional exam of Pharmacology

**Sample selection**: Academically poor performing students were selected from the mark list of first sessional exam. (All students who got less than 50 % marks). The students were randomized into two groups using random number table. One group received academic support alone. The second group received academic support, sessions on study skills, stress-coping strategies and counselling regarding their academic and non-academic problems.

**Inclusion Criteria:** Students who got less than 50% marks and willing to give informed consent

**Study tools:** A questionnaire was used to collect the basic socio demographic information, habits and other factors that can affect academic performance. *Study skills self-assessment* inventory was used to assess the study skills of participants and design the remedial sessions. A pre-test and a post-test questionnaire was used to assess effectiveness of educational intervention

**Intervention:** The selected students were given remedial teaching (Specialized instruction for students deviating from the expected norm.)

One group (26 students) received academic support alone. The second group (27 students) received academic support, sessions on study skills, stress-coping strategies and counselling regarding their academic and non-academic problems.

### Data collection process

After obtaining informed consent, all the second MBBS students were given a questionnaire enquiring the basic socio demographic information, habits and other factors that can affect academic performance. This was followed by administration of *Study skills self-assessment inventory* to assess the study skills of participants and aid in effectively designing the remedial sessions. Then from first sessional exam results in Pharmacology, all students who got less than 50% marks and who were willing to participate in the study was enrolled for remedial teaching. An informal discussion was held with the learners. After taking an informed consent from these students, a pre-test (MCQs& SAQs) on two selected Pharmacology topics was conducted one week before the remedial sessions.

The students were randomized into two groups using random number table. One group received academic support alone. The academic support was given after regular class timings and included providing study materials on the two selected pharmacology topics and small group discussions. For the other group, along with academic support, a session on study skills was conducted with emphasis on listening and taking notes in class, time management, study practices, concentration, recall, test anxiety and method of answering questions. In the following week, the second group of students were counselled regarding improvement of academic performance. The students were also introduced to various stress-coping strategies. Post interventional assessment of both the groups was done again by MCQs& SAQs. A post-programme questionnaire was administered to the students who participated in the program to assess whether the intervention was of any benefit. The second group was also given study skills& stress management sessions after conclusion of the study.

## Data analysis

The data was entered in MS Excel and was analysed using the SPSS software. Associations were tested using Chi square test. Mann–Whitney U test was used to compare scores between intervention and control group and Wilcoxon signed-rank test was used to compare pre- and post-intervention performance of the students who participated in the program.

## RESULTS

Seventy-eight students participated in the study by completing the *study skills self-assessment inventory. Study skills self-assessment inventory* was administered to assess the study skills of participants and aid in effectively designing the remedial sessions. Out of 78 students who completed the inventory, 33 (42.3%)were males and 45 (57.7%) were females. Only 25 students (32.1%) out of 78 participating students had passed first sessional pharmacology examination. It was observed that the pass percentage is more among females and this observation is statistically significant. (Chi-Square value 10.43, df – 1, p value 0.001)

No association was observed between having a regular hobby and academic performance. Association between smoking(cigarettes) habits and academic performance could not be elicited as all the participants were non-smokers. No association was observed between academic performance and consumption of caffeine containing beverages or social life outside the college or number of hours spend on extracurricular activities.

Though not statistically significant, better academic performance was observed in participants who slept for optimum hours (> 6hours/day or < 8 hours/day). It was observed that studying alone or with a colleague or in groups has no statistically significant effect on academic performance. English proficiency also was observed to have no significance effect on exam performance. Majority of the students gave a response of not studying pharmacology on a daily basis.

34.6% of the students gave the opinion that personal attitude was the most significant factor which improved their learning. Interestingly 71% of students gave personal attitude and learning patterns as the most important factor adversely affecting their academic performance. 45.5% of participants gave the opinion t^6^hat most of the time, ^6^they were motivated to participate in the class.

<u>The following results were obtained on application of *study skills self-assessment inventory* on the participants.(</u> Table1)

**Table1:**
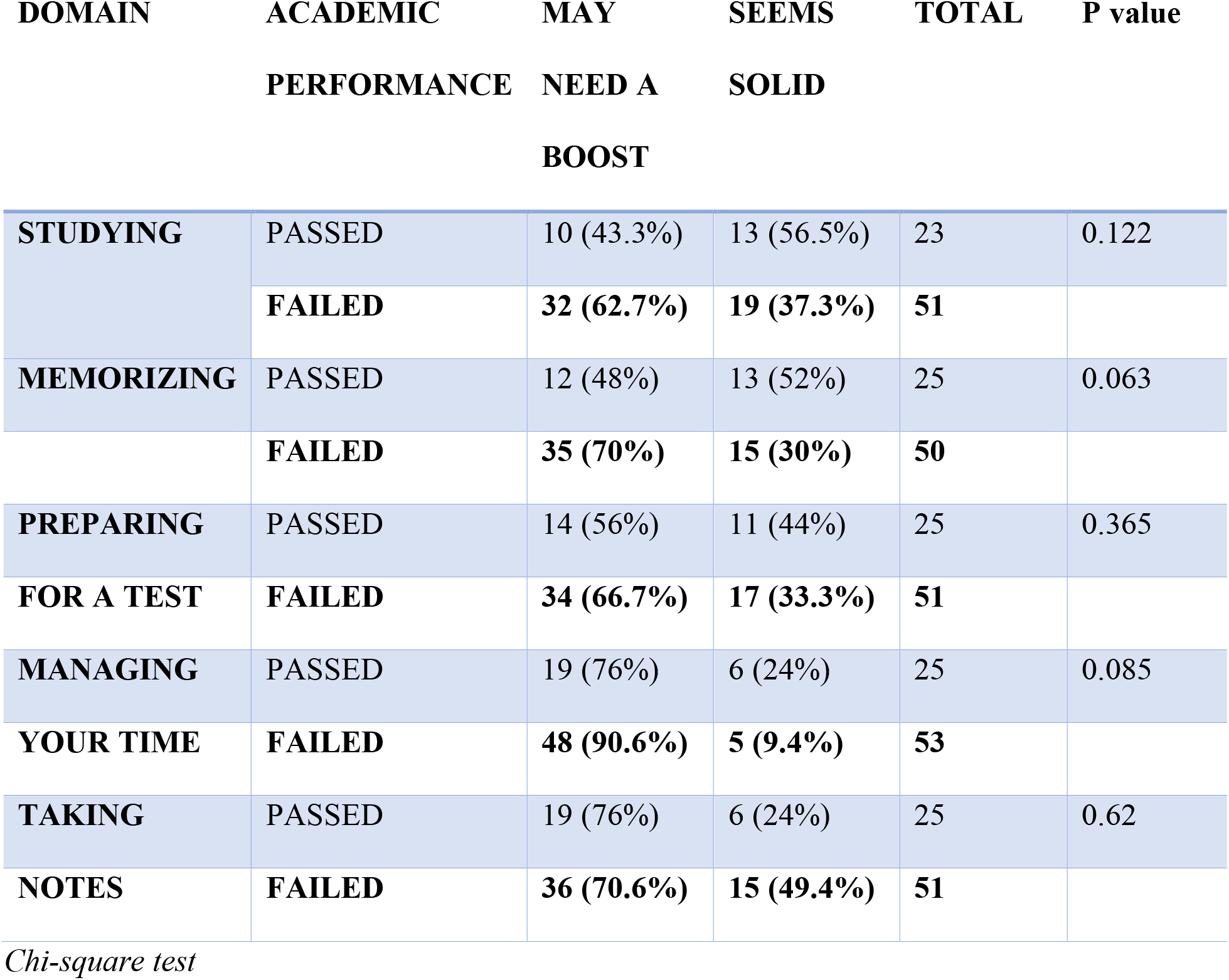
Domains under study skills inventory Vs. exam performance.

| **DOMAIN** | **ACADEMIC**<br>**PERFORMANCE** | **MAY**<br>**NEED A**<br>**BOOST** | **SEEMS**<br>**SOLID** | **TOTAL** | **P value** |
| --- | --- | --- | --- | --- | --- |
| **STUDYING** | **PASSED** | 10 (43.3%) | 13 (56.5%) | 23 | 0.122 |
|  | **FAILED** | **32 (62.7%)** | **19 (37.3%)** | **51** |  |
| **MEMORIZING** | **PASSED** | 12 (48%) | 13 (52%) | 25 | 0.063 |
|  | **FAILED** | **35 (70%)** | **15 (30%)** | **50** |  |
| **PREPARING**<br>**FOR A TEST** | **PASSED** | 14 (56%) | 11 (44%) | 25 | 0.365 |
|  | **FAILED** | **34 (66.7%)** | **17 (33.3%)** | **51** |  |
| **MANAGING**<br>**YOUR TIME** | **PASSED** | 19 (76%) | 6 (24%) | 25 | 0.085 |
|  | **FAILED** | **48 (90.6%)** | **5 (9.4%)** | **53** |  |
| **TAKING**<br>**NOTES** | **PASSED** | 19 (76%) | 6 (24%) | 25 | 0.62 |
|  | **FAILED** | **36 (70.6%)** | **15 (49.4%)** | **51** |  |

*Chi-square test*

The different domains under the inventory are *studying, memorizing, preparing for test, managing time and taking notes*. Each domain has 5 sub questions under it. For each domain, maximum score is 50 and minimum zero. These scores are divided into two.

A score of 35 to 50 means the skills **“seems solid”**. A score 0 to 34 means skills may **“need a boost”**.

While looking at participant’s domain-wise score, following median scores were obtained.

- Studying-30
- Memorizing- 25
- Preparing for a test-30
- Managing your time-15
- Taking notes-27.5

According to this inventory, majority of our students fall far below the recommended cut off for skills. (Table1) The difference observed in skills in the domain ***“studying*”** across passed and failed students in first sessional exams in pharmacology, is statistically significant. p 0.012. (Mann-Whitney U)

The difference observed in skills in the domain ***“managing your time*”** across passed and failed students in first sessional exams in pharmacology, is statistically significant. p 0.005. (Mann-Whitney U)

While analyzing the different domains under study skills self-assessment inventory using the recommended cut off, for all participants,

- 42(56.8%) *may need a boost* under the domain “studying”.
- 47(62.7%) *may need a boost* under the domain “memorizing”.
- 48(63.2%) *may need a boost* under the domain “preparing for a test”.
- 67(85.9%) *may need a boost* under the domain “managing your time”.
- 55(72.4%) *may need a boost* under the domain “taking notes”.

Though not significant, overall this inventory has identified that all those who failed in the exam “may need a higher boost” in all the domains when compared to those who passed.

The inventory has identified that female participants as “seems solid” in all the domains when compared to male participants. This difference is statistically significant in the domain memorizing.

## Remedial sessions

Remedial teaching was given to all who failed in Pharmacology in First Sessional Exam (53 students). 29 male and 24 female participants attended the remedial sessions. The participants were divided into two and one group was given “academic support alone” and the other group was given “academic support and specialized instructions”. Their baseline characteristics were similar. (First sessional pharmacology exam marks, pre-test marks). There is no significant difference in distribution of marks in pharmacology first sessional exam in both intervention groups.

The improvement in test scores as assessed by a post-test among participants of remedial sessions was statistically significant.(Figure 1) Though there is an observed difference in marks gained between two intervention groups, the difference is not statistically significant. (T test, P value 0.16)

**Figure 1.**
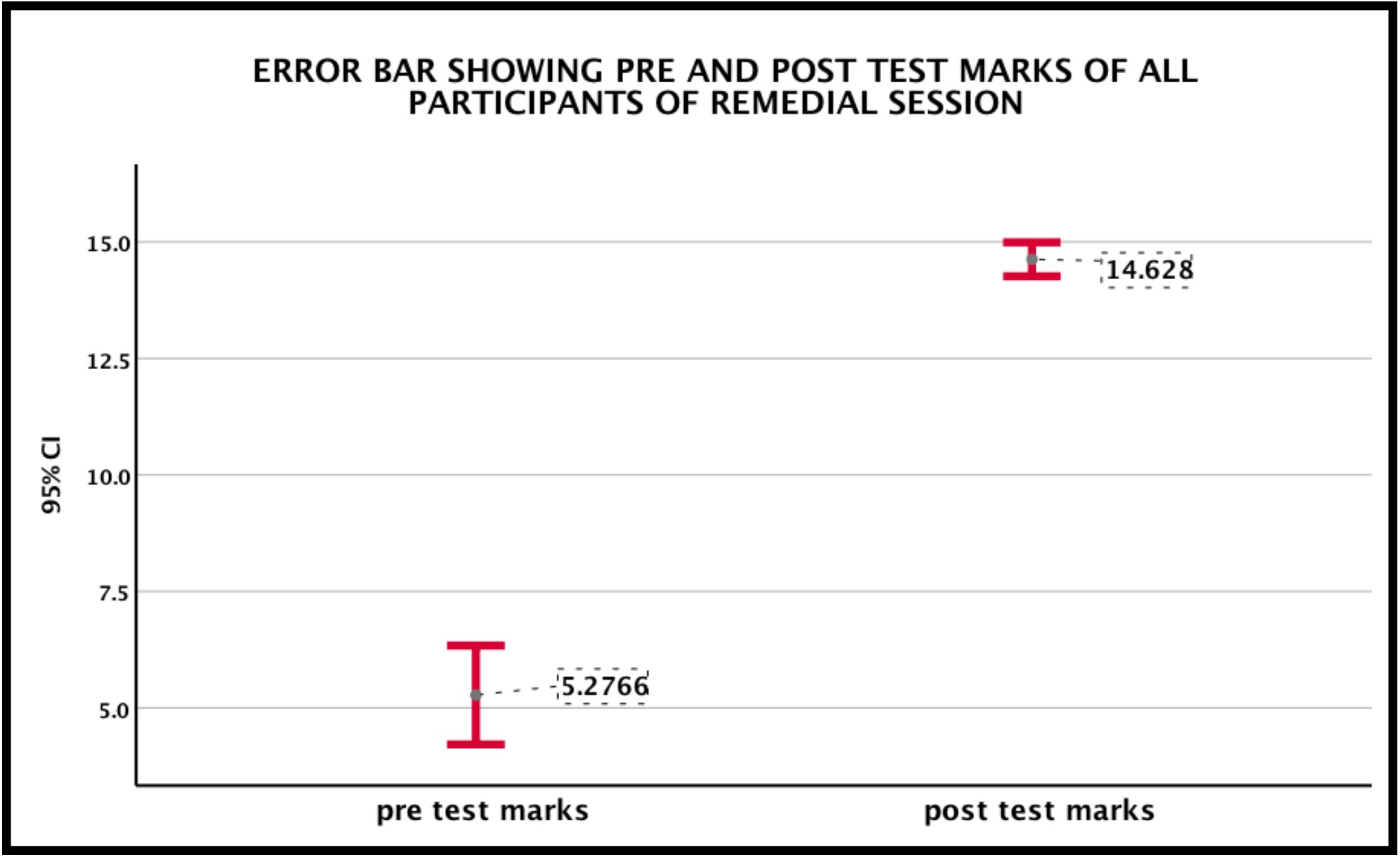
Error bar showing pre and post test marks of all participants of remedial session

## Post program questionnaire

A post-programme questionnaire was given to the students who participated in the program to assess whether the intervention was of any benefit. 49.1% of students who attended the remedial teaching sessions strongly agreed that that the sessions helped them to understand the topics better.

The participants agreed that the sessions 1) motivated them to learn *2*) improved their interest in the subject *3*) they would like to use this method for learning more topics *4*) improved their level of confidence *5*) they put effort into learning the material covered in this intervention and *6*) they were challenged to do their best work.

The majority of students also believed that they were able to better understand the drugs, easily able to grasp the topic & its contents, study the topics thoroughly, recollect more things and thus improve confidence, could revise again &again and the sessions helped to focus on topics which were most important. The students also commented that this was a very good method to learn pharmacology because of the interactive sessions, creative slides, repetition of important points of the topic and thus very effective in remembering contents of the topic.

When asked what aspects of the sessions were most beneficial, the most common response was interactive sessions, followed by responses such as creative slides, diagrammatic representations, schematic diagrams of mechanism of action &flow charts, simplification of topic, focus on important points, pre-test followed by discussions and post-test, repetition of important points, presentation of the teacher, repeated questions asked by the teacher and they were also able to discuss doubts and solve them. When asked what they did not like about the sessions, the students did not find any.

When asked for suggestions to improve teaching in Pharmacology, the suggestions received were to include this type of interactive learning with pictures and animations in teaching the subject, so as to give a better understanding of topic.(Figure 2)

**Figure 2.**
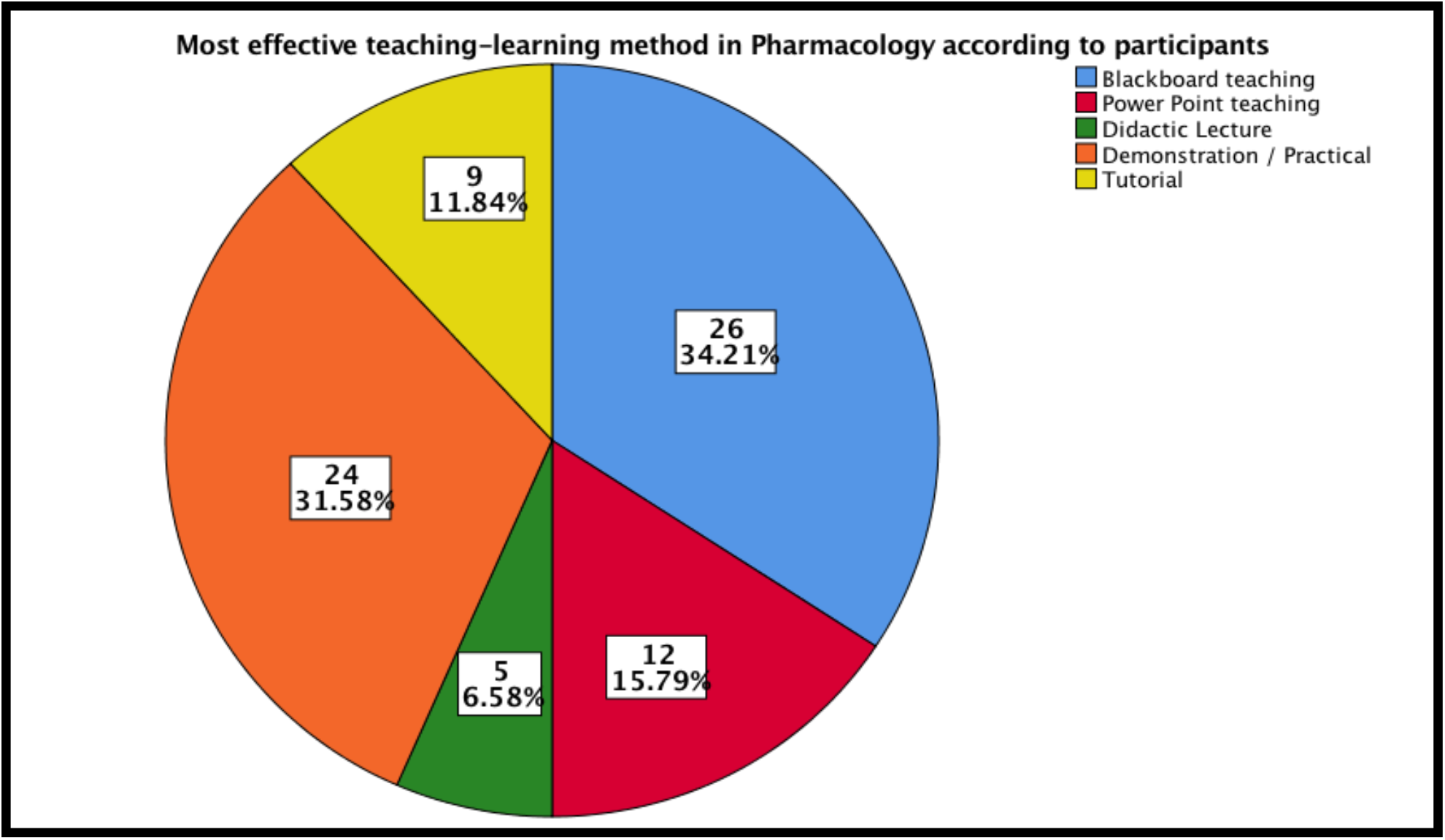
Most effective teaching-learning method in Pharmacology according to participants

## DISCUSSION

Literature establishes that the demands of a medical school is difficult to cope with. About 10-15% of medical students face substantial academic difficulties during their course.^12,13^ Hence, early and timely intervention in the form of targeted remediation is warranted for students experiencing academic difficulties. Even though evidence shows that targeted remediation can benefit such students, little is known about the best approach for remediation in medical students.^14^

This study provides evidence that targeted remediation after assessing the deficiencies in study skills of the students, can substantially improve academic performance of poorly performing medical students. The study emphasises the importance of identification of learning problems at an early stage and intervention in the form of adequate remediation to such students. ^15^

A remedial teaching intervention should always be learner-centric, tailored to an individual student’s needs. This is because the type of support a student needs can vary, with some students just needing assistance in time management skills or improvement in study techniques whereas other students might need more than one strategy, possibly including enhanced learning support throughout their academic years.^15^

This variability necessitates that every remedial intervention should start with a diagnosis of the individual learner’s needs thus enabling teachers to provide individualised remediation to the student.^16^ This attempt at diagnosis is always better done by teachers who are currently teaching the student. The remediation sessions should ideally include multiple, spaced conversations with the student probing into the reasons and attempting an exact identification of remedial measures the student needs. ^12,13^ The current study has attempted to achieve the pedagogical diagnosis by two steps. In the first step, we utilised a questionnaire to get a surface knowledge of the sociodemographic background of all the students and his/her study skills, time management skills etc. by a self-assessment questionnaire.^17^ This was followed by further interaction with the study group, attempting to get an in-depth knowledge about the reasons for his/ her poor academic performance. Then remedial sessions were designed according to the perceived needs of the students. ^15^

Remedial interventions must undergo continuous and rigorous evaluation throughout the academic programme. The interventions should be facilitated by teachers capable of pedagogical diagnosis who can also play the role of an encouraging mentor. They should ideally be role models for the students and should possess high degrees of teaching presence and practical wisdom.^18^ This is necessary because significance differences in remediation outcomes have been observed with experienced and inexperienced teachers.^18^ The success of remedial interventions can also be enhanced when conducted in small groups, by experienced teachers who are familiar with the content and process. ^18^

## CONCLUSIONS

The study proves that remediation measures enabled by simple tools like a study skills inventory can help identify and correct learning difficulties of medical students at an early stage. Such tailored or individualised remediation measures can greatly enhance the academic performance of undergraduate medical students and help them make satisfactory progress on the course.

### Limitations of this study

1. The findings of this study may not be generalizable and may be context specific, specific to this cohort or specific for learning pharmacology.
2. Small sample size
3. Even though the *Study skills self-assessment inventory* is valid and internally consistent, it is a self-reporting instrument and thus the true approach to learning of students may not be reflected, especially if they answered the questions in a way that they thought would have been the ‘expected answers’.
4. Examination of only a small range of demographic variables

## Data Availability

Not applicable

## Acknowledgements

The author is grateful to all the participants who have participated and provided feedback for completion of the study. The research was supported by MCI Nodal Centre for Faculty development, Government Medical College, Kottayam, Kerala, India.

## Ethical considerations

Institutional Research Committee and Ethics Committee clearance was obtained from Institutional Human Ethics Committee, Government Medical College, Parippally, Kollam, Kerala, India. IEC.No.3/EC-3/2019/GMCKLM Dated 30/07/2019

Permission of Institutional Head was obtained.

Informed consent was obtained from participating students.

## Disclaimers

None

## Previous presentations

None

